# Are current guidelines overcautious regarding refeeding of patients with severe anorexia nervosa: a retrospective cohort study

**DOI:** 10.1101/2020.04.26.20050799

**Authors:** Oana Ciobanasu, Bhavisha Seth, Iryna Terekh, Alessandro Bruno, Agnes Ayton

## Abstract

Weight restoration is an integral part of managing anorexia nervosa patients and has been found to be associated with electrolyte and fluid abnormalities gathered under the umbrella term refeeding syndrome, which has led to cautious initiation of caloric regimes. This study looks at how a sample of severe anorexia nervosa patients were managed using higher rates of refeeding than the ones currently recommended.

**Design:** retrospective cohort study of consecutive patients with severe eating disorders admitted to an UK specialist tertiary centre. The treatment programme uses a weight restoration regime starting at 1000 kcal/day, increased to 1500 kcal/day after two days and to 2000 kcal/day after seven days. The main outcome was the rate of hypophosphatemia, hypokalemia and hypomagnesemia within the first two weeks of weight restoration. The secondary outcomes included rate of weight gain and tendency of electrolyte shift.

**Results:** 83% of the patient sample were categorised as extreme anorexia nervosa (BMI <15). 11.3% of patients developed hypophosphatemia, 11.3% had hypomagnesemia, 42% had hypokalaemia. The lowest levels were found between the 5-6 days after starting refeeding. The vast majority of electrolyte abnormalities fell into the mild category. Electrolyte abnormalities were easily corrected by oral supplementation, and only 5% required iv replacement. The only significant predictor for hypophosphatemia was a BMI below 13.

**Conclusions:** The majority of extremely ill patients with anorexia nervosa tolerate refeeding starting at 25-30kcal/kg. Oral supplementation is effective, so overcautious refeeding is unnecessary, provided that the patient is carefully monitored.

## Introduction

Outside specialist services, eating disorders are a poorly understood group of diagnoses, with limited knowledge of the presentation, risks and management. This is documented not only in primary care physicians(1) but also in hospital doctors including general psychiatrists(2-4). This is concerning as the diagnosis, as well as the treatment and management of people with eating disorders is likely to be initiated outside specialist services. In emergency situations, the care of anorexia nervosa patients may take place in an acute medical setting where the majority of staff is unlikely to have the adequate training in managing such patients. As a result of the overall poor understanding of eating disorders in practice there is heavy reliance on the use of existing guidance. Recognising this problem, in 2010 the Royal College of Psychiatrists in collaboration with other stakeholders, including the Royal College of Physicians, established the ‘Management of really sick patients with anorexia nervosa’ (MARSIPAN) group and developed a consensus report to prevent patients with severe anorexia nervosa dying in hospitals(5). One of the issues that were discussed by the MARSIPAN group was that some patients were doing worse than expected following admission to acute hospitals due to over-cautious re-feeding regimes. We can speculate that the reasons behind such regimes are multiple and not well researched, but it is clear that one of them is concern over emergence of refeeding syndrome in these patients. This is reflected in the MARSIPAN recommendations which suggest starting refeeding with 5-10kcal/kg/day. This was a result of consensus in the absence of sufficient evidence and was based on the 2006 NICE guidelines regarding nutrition support for adults, even though the NICE guidelines did not cover malnutrition in anorexia nervosa and related disorders(6). These recommend refeeding rates of only 5 kcal/kg/day in extreme cases (for example, BMI less than 14 kg/m2 or negligible intake for more than 15 days). The NICE guidelines were reviewed in 2017 with no changes to the recommendations due to insufficient new evidence.

Refeeding syndrome is potentially fatal, and can present with cardiac, pulmonary and neurological symptoms and is more prevalent in the initial stages of weight restoration(7). The hallmark of re-feeding syndrome is hypophosphatemia, triggered by the shift from fat and protein to carbohydrate metabolism which stimulates the previously decreased secretion of insulin and leads to intracellular uptake of phosphate and thiamine. While low phosphate is the most frequent indicator of re-feeding syndrome, other fluid and electrolyte imbalances may be present, such as hypokalaemia and hypomagnesemia.

The MARSIPAN guideline advises a starting calorie regime of 20 kcal/kg/day in specialist eating disorder settings and 5-10 kcal/kg/day in medical in-patient settings, especially in the presence of severity indicators, such as very low initial weight (BMI <12), electrolyte abnormalities at baseline, significant ECG abnormalities, etc(5). However, US and Australian guidelines are less cautious, but the evidence base remains limited(8).

This present study analyses the frequency of re-feeding syndrome in a sample of patients with extreme and severe anorexia nervosa who are initiated on caloric regimes exceeding the recommended values of the current guidelines.

## Methods

This was a retrospective cohort study including all patients admitted with a severe or extreme anorexia nervosa or avoidant restrictive food intake disorder (ARFID) as defined by DSM-5 (severe: BMI 15-15.99; extreme: BMI<15) between January 2014 and December 2016 (n= 71) to a tertiary eating disorder centre in Oxford, United Kingdom. The main inclusion criteria were a Body Mass Index (BMI)<16 and a diagnosis of anorexia nervosa. There were no exclusions from the study population. Demographic and clinical information was manually collected using the electronic records. The local biochemistry laboratory was contacted and asked to provide the electronic biochemistry results for the patients. The project protocol was approved by the Oxford Health NHS Foundation Trust Audit’s department, and recorded as a clinical audit that did not require ethics approval. It did not require informed patient consent due to the retrospective design and anonymization of patient identifiers.

The descriptive variables recorded for each patient included: age, gender, DSM-5 diagnosis, weight and BMI on admission. The diagnoses (DSM 5) of all admissions were categorised into three main subtypes – restrictive, binge-purge and ARFID. The main outcome measure was the development of refeeding syndrome, defined as the presence of hypophosphatemia, hypokalaemia or hypomagnesemia at any point during the first two weeks. Hypokalaemia was defined as serum potassium levels of less than 3.5 mmol/L, for serum phosphate and magnesium the abnormal levels was categorised as levels below 0.7mmol/L, and below 0.68 mmol/L respectively. All electrolyte abnormalities were further divided according to severity in three categories: mild, moderate and severe, the threshold of each are given in Table 2. The lowest values of all three serum electrolytes during the first two weeks were recorded.

**Table 1.**
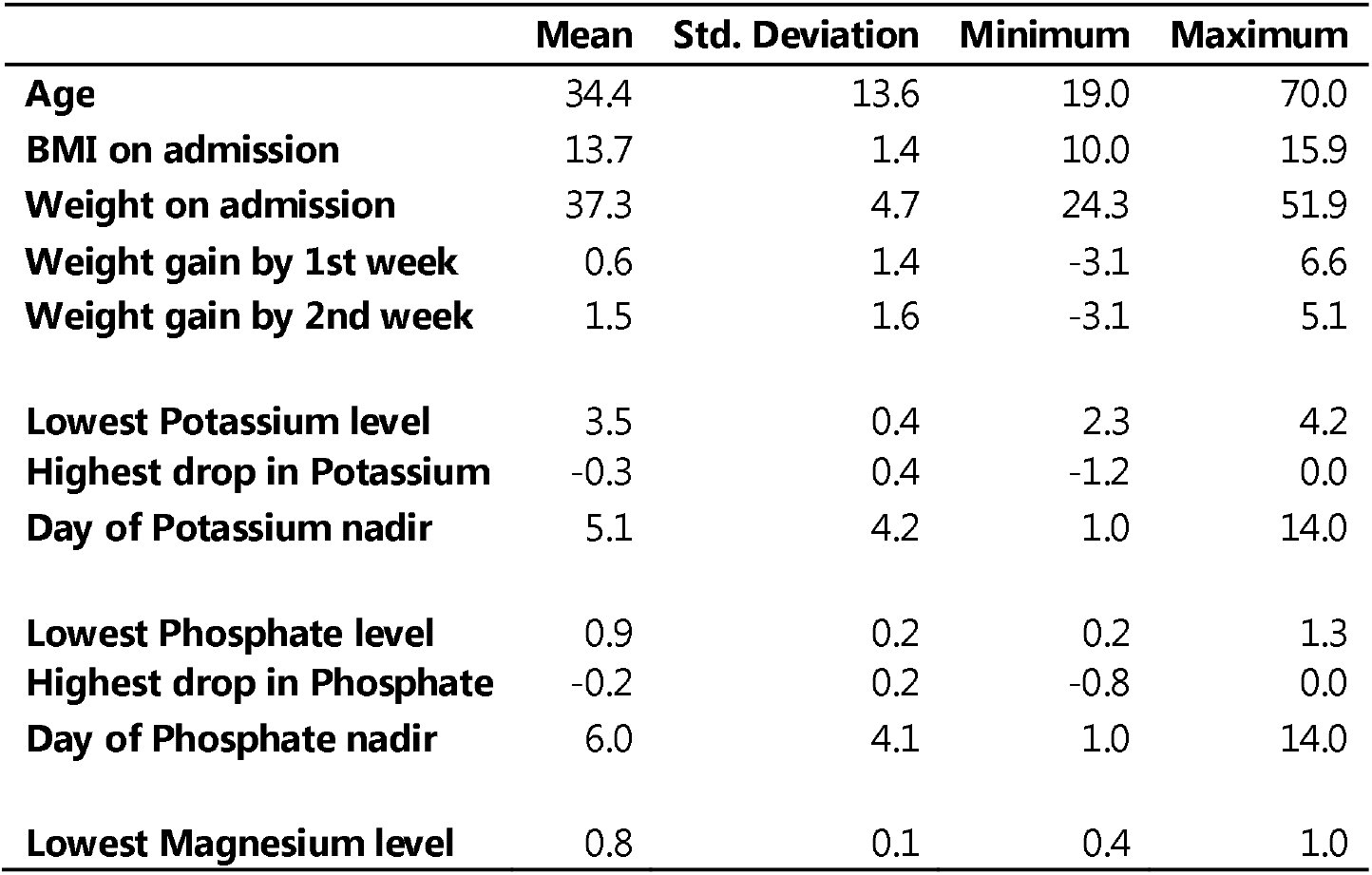
Descriptive statistics

|  | Mean | Std. Deviation | Minimum | Maximum |
| --- | --- | --- | --- | --- |
| **Age** | 34.4 | 13.6 | 19.0 | 70.0 |
| **BMI on admission** | 13.7 | 1.4 | 10.0 | 15.9 |
| **Weight on admission** | 37.3 | 4.7 | 24.3 | 51.9 |
| **Weight gain by 1st week** | 0.6 | 1.4 | -3.1 | 6.6 |
| **Weight gain by 2nd week** | 1.5 | 1.6 | -3.1 | 5.1 |
| **Lowest Potassium level** | 3.5 | 0.4 | 2.3 | 4.2 |
| **Highest drop in Potassium** | -0.3 | 0.4 | -1.2 | 0.0 |
| **Day of Potassium nadir** | 5.1 | 4.2 | 1.0 | 14.0 |
| **Lowest Phosphate level** | 0.9 | 0.2 | 0.2 | 1.3 |
| **Highest drop in Phosphate** | -0.2 | 0.2 | -0.8 | 0.0 |
| **Day of Phosphate nadir** | 6.0 | 4.1 | 1.0 | 14.0 |
| **Lowest Magnesium level** | 0.8 | 0.1 | 0.4 | 1.0 |

**Table 2.** Table 2. Electrolyte abnormalities by diagnostic sub-category

| Electrolyte | AN – BP (n = 16) | AN - R (n = 53) | Total (n = 71) |
| --- | --- | --- | --- |
| **Phosphate plasma levels** |  |  |  |
| ❖ Normal (0.7 – 1.4 mmol/L) | 16 (100%) | 45 (85%) | 63(89%) |
| ❖ Hypophosphatemia | 0 (0%) | 8 (15%) | 8 (11%) |
| • Mild (0.6 – 0.69 mmol/L) | 0 (0%) | 2 (4%) | 2 (3%) |
| • Moderate (0.3 – 0.59 mmol/L) | 0 (0%) | 5 (9%) | 5 (7%) |
| • Severe (<0.3 mmol/L) | 0 (0%) | 1 (2%) | 1 (1%) |
| **Magnesium plasma levels** |  |  |  |
| ❖ Normal (0.7 – 1 mmol/L) | 13 (81%) | 48 (91%) | 63 (89%) |
| ❖ Hypomagnesemia | 3 (19%) | 5 (9%) | 8 (11%) |
| • Mild (0.5 – 0.7 mmol/L) | 2 (12.5%) | 4 (7%) | 6 (8%) |
| • Moderate (0.4 – 0.5 mmol/L) | 1 (6.5%) | 1 (2%) | 2 (3%) |
| • Severe (<0.4 mmol/L) | 0 (0%) | 0 (0%) | 0 (0%) |
| **Potassium plasma levels** |  |  |  |
| ❖ Normal (3.5 – 5.5 mmol/L) | 6 (37.5%) | 34 (64%) | 41 (58%) |
| ❖ Hypokalaemia | 10 (62.5%) | 19 (36%) | 30 (42%) |
| • Mild (3.0-3.5 mmol/L) | 4 (25%) | 16 (32%) | 20 (28%) |
| • Moderate (2.5 – 3.0 mmol/L) | 5 (31%) | 3 (9%) | 9 (13%) |
| • Severe (<2.5 mmol/L) | 1 (6%) | 0 (2%) | 1 (1%) |

Secondary outcomes included: the weight gain and BMI after one and after two weeks of treatment and transfer to a general hospital for intravenous electrolyte supplementation at any point during the first two weeks of admission. We also measured the tendency of electrolyte shift, calculated by subtracting the lowest serum level from the admission serum level for all three electrolytes. We also analysed whether there was a significant difference in outcomes in between different BMI and diagnostic subgroups. These variables were entered into an electronic spreadsheet and collated for analysis.

As summarised in Figure 1, the standardised nutritional provision for patients admitted to Cotswold House begins at 1000 kilocalories on day one and this is subsequently increased to 1500 kilocalories by day three and to 2000 kilocalories after seven days. They undergo regular blood testing, the frequency of which is influenced by clinical judgement based on biochemical results, but as a minimum, they will have at least three blood tests during the first week. Oral phosphate, potassium and magnesium supplements are given to patients with electrolyte levels below or just above the laboratory cut-off for normal levels.

**Figure 1.**
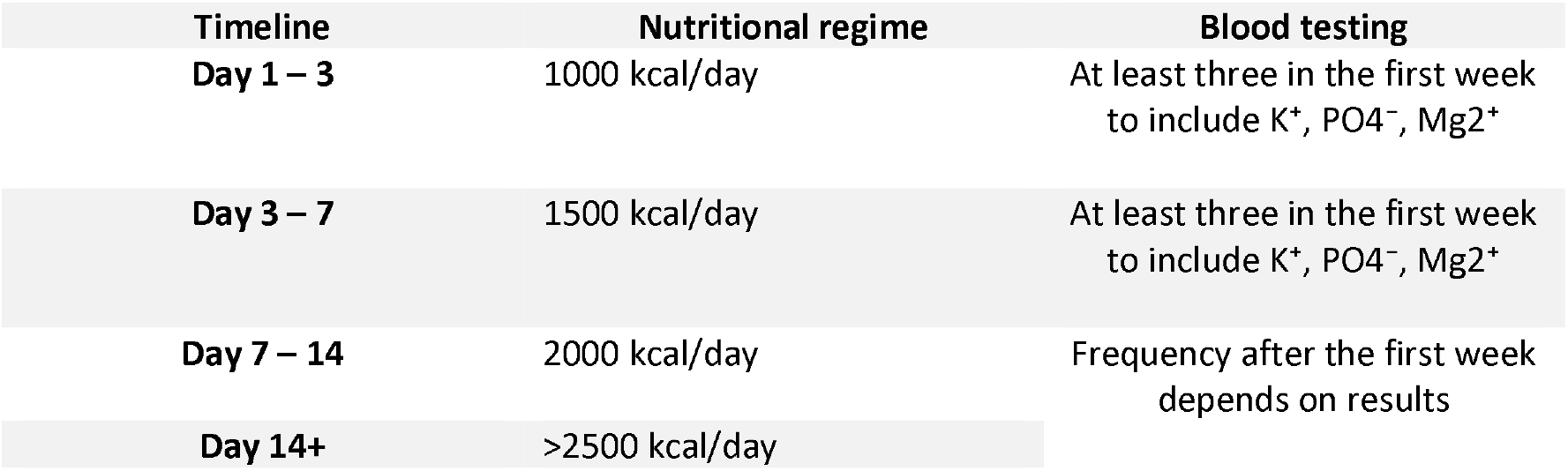
Refeeding regime at Cotswold House.

Statistical analysis was performed using SPSS 22. Descriptive statistics were calculated for the entire study population. Group means, standard deviations and ranges are reported for demographic and clinical data. Pearson’s coefficients were used to calculate correlation. Chi squared was used for categorical data. Paired t-tests, student t-tests and one-way variance of analysis were used to assess variation for continuous variables.

## RESULTS

71 admissions met the inclusion criteria and were included for analysis. 53 had restrictive anorexia, 16 had the binge-purge subtype and 2 ARFID. There were three male and 68 female patients in the sample. 83% of the study population had a BMI of less than 15, which using DSM-5 definition falls into the extreme severity category. 31% of the sample (n = 21) had a BMI of less than 13 which according to MARSIPAN guidelines is the high risk of physical deterioration category. There were no statistically significant differences between the restrictive and binge-purge subgroups in terms of admission weight, BMI and age.

Table 1 summarises age, BMI and weight on admission and rates of weight gain throughout the first two weeks. It also includes the recorded changes in potassium, phosphate and magnesium levels. The refeeding regimen (as per Figure 1) started with a mean 26.7kcal/kg/day on day 1-2 (range 20-41), followed by 40 kcal/kg/day on day 3-7, and 53.5 kcal/kg/day on day 7-14, and was adjusted afterwards depending on weight restoration. The overall incidence of hypokalaemia was 42%, hypophosphatemia was 11%, and hypomagnesaemia was 11%. Tables 2 and 3 detail the recorded frequencies for the total sample and for diagnostic and BMI sub-groups. In all, except one case, oral supplementation was sufficient to correct abnormalities. The abnormalities occurred within the first two weeks; the mean nadir was on days 5-6. BMI and diagnostic subcategory did not significantly impact on the rate of weight gain. There was a statistically significant moderate correlation between potassium levels and DSM-V diagnosis (r(67) = -.388, p <.001), indicating that a diagnosis of binge-purge anorexia was overall correlated with lower levels of potassium, albeit there was no statistically significant difference in hypokalaemia incidence between the groups: *X*^*2*^ (1, N = 69) = 3.583, p = .0583. There were no recorded cases of hypophosphatemia in the binge-purge subgroup. BMI was positively correlated with phosphate level, r(69) = .340, p <0.005 but not with potassium or magnesium levels. Patients with a BMI lower than 13 were more likely to develop hypophosphatemia, with an OR of 24.5, 95% confidence interval (2.7 to 216), p=0.004. There was no statistically significant difference in the odds of developing hypokalaemia or hypomagnesemia between the different BMI groups. Overall the patients who developed hypophosphatemia had a significantly lower mean admission BMI compared to those who did not: 11.9 vs 13.8, t(69) = -4.1, p < .0001.

**Table 3.** Electrolyte abnormalities by BMI category

| Electrolyte | BMI <13 (n = 21) | BMI 13 – 15 (n= 36 ) | BMI > 15 ( n= 14) |
| --- | --- | --- | --- |
| **Phosphate plasma levels** |  |  |  |
| ❖ Normal (0.7 – 1.4 mmol/L) | 14 (67%) | 35 (97%) | 14 (100%) |
| ❖ Hypophosphatemia | 7 (33%) | 1 (3%) | 0 (0%) |
| • Mild (0.6 – 0.69 mmol/L) | 2 (9.5%) | 0 (0%) | 0 (0%) |
| • Moderate (0.3 – 0.59 mmol/L) | 4 (19%) | 1 (3%) | 0 (0%) |
| • Severe (<0.3 mmol/L) | 1 (5%) | 0 (0%) | 0 (0%) |
| **Magnesium plasma levels** |  |  |  |
| ❖ Normal (0.7 – 1 mmol/L) | 19 (90%) | 31 (86%) | 13 (93%) |
| ❖ Hypomagnesemia | 2 (10%) | 5 (14%) | 1 (7%) |
| • Mild (0.5 – 0.7 mmol/L) | 2 (10%) | 3 (8%) | 0(0%) |
| • Moderate (0.4 – 0.5 mmol/L) | 0 (0%) | 2 (6%) | 1(7%) |
| • Severe (<0.4 mmol/L) | 0 (0%) | 0 (0%) | 0(0%) |
| **Potassium plasma levels** |  |  |  |
| ❖ Normal (3.5 – 5.5 mmol/L) | 11 (52%) | 21 (58%) | 9 (64%) |
| ❖ Hypokalaemia | 10 (48%) | 15 (42%) | 5 (36%) |
| • Mild (3.0-3.5 mmol/L) | 7 (33%) | 11(31%) | 2 (14%) |
| • Moderate (2.5 – 3.0 mmol/L) | 3 (14%) | 3 (8%) | 3 (21%) |
| • Severe (<2.5 mmol/L) | 0 (0%) | 1 (3%) | 0 (0%) |

## Discussion

The current study shows that initiating refeeding at over 1000 calories per day and rapidly increasing the intake to 2000 kcal by day seven in a sample of patients with severe or extreme anorexia nervosa has a low frequency of severe refeeding complications: in our sample only one patient had phosphate levels drop below 0.3 mmol/L and only one other patient had their potassium levels drop below 2.5 mmol/L. Furthermore, only one patient required IV supplementation, the rest were managed on the specialist eating disorder ward through oral supplementation. These results indicate that even patients with extreme malnutrition can be safely managed with a refeeding meal plan starting between 20-40kcal/kg/day if electrolytes are carefully monitored and supplemented as required. This is significantly higher than the current MARSIPAN or NICE recommendations and indicates excessive cautious is not needed, as previously suggested. A higher calorie regime, such as the one used in our study, will likely lead to faster weight gain and reduce overall length of stay. It is also worth noting the unit in which the study was conducted is operating under a rigorous regime which includes regular reviews of biochemical findings, blood tests and patient examinations, which likely contributed significantly to the reduced incidence of severe electrolyte disturbances and lack of fatal outcomes. Our findings also show that, as expected, patients with a history of binge-purge type of anorexia have lower potassium levels, and patients below a BMI of 13 are more likely to develop hypophosphatemia, however of note the majority of reported cases fell in the mild severity category.

Since the 2^nd^ revision of the MARSIPAN guidelines, and the NICE guidelines, there has been a growing body of evidence that looks at patients with anorexia nervosa and the directions to treatment. A 2016 review(9) identifies 27 studies focusing on approaches to refeeding in adults and adolescents with anorexia nervosa, some of which have refeeding syndrome as an outcome. A further review published the same year(7) looks specifically at the rates of refeeding syndrome and identifies 16 studies focusing on anorexic samples. A more recent one, published in 2019(10) specifically looks at the refeeding process of patients with anorexia nervosa, focusing on clinical outcomes and identifies 19 papers. Of all the studies identified by the three reviews only six(11-16) adult sample studies use a higher calorie approach, delivered through oral nutrition, similar to our study and report rates of refeeding syndrome. Most of the samples are small and include a wide range of BMI values and only one of these studies(12) report rates of hypokalaemia and hyponatraemia. The rates of hypophosphatemia reported by the other studies range from 6.5%(13) to 45%(15). BMI and starting calorie regime are comparable in these studies, however there is significant variation in the variables reported, and as a result it is difficult to establish whether the rate difference is protocol or sample characteristics related, or a mixture of both.

One of the strengths of this study is the large sample size, including mostly patients presenting with extremely severe anorexia nervosa – these are the types of patients likely to be admitted to a medical setting for initial refeeding and therefore likely to be receiving a lower caloric intake which may lead to slower weight gain(15). Additionally, we have included consecutive patients admitted to a tertiary specialist service over a period of three years therefore providing good external validity to the study. Of note, while other studies(12) have reported the BMI of patients with the binging-purging subtype of AN to be significantly higher than that of the restrictive group, our study found both groups had similar average BMI: 13.8 and 13.6. Similar to previous studies(14) we found that the only significant predictor of hypophosphatemia was lower admission BMI.

In terms of limitations, this is a retrospective cohort study and not a randomised controlled trial. As in other studies, we have given patients supplements prophylactically, when blood results indicated phosphate, magnesium and phosphate levels to be close to the low range of normal. We believe that while this may result in an underestimation of the rate of refeeding syndrome, it’s also reflective of patient-safety orientated practice and not doing so would not be ethically defensible in clinical research. In addition, we recognize that for the purpose of this study our definition of refeeding syndrome does not take into consideration clinical symptoms and is based solely on biochemical values. This aspect, together with the conclusions drawn from reviewing similar studies, highlights the fact that a unified definition of the refeeding syndrome is not yet available and outlines the need for such, as it would allow for more research homogeneity and more robust conclusions to be drawn from the body of evidence available. In addition, given the fact that most available evidence is based on relatively small samples, we highlight the need for further multi-center standardized research which would allow for robust and relevant guidance to be implemented.

## Conclusions and recommendations

Current available research on refeeding syndrome in adults is limited, but several studies, including our own, suggest available guidelines may be too cautious. Randomised controlled trials in adolescent samples show that refeeding at a rate of 20-30 kcal/kg/day is safe and similar studies are needed in adult samples.

## Data Availability

Pertinent clinical data is included in the body of the paper, further anonymized study data can be provided upon request.

## Acknowledgements

The authors would like to thank Dr Brian Shine (Consultant chemical pathologist at Oxford University NHS Foundation Trust) for his contribution to obtaining the data. The authors would also like to thank Julie Lambert and Lucy Gardner (Dieticians at Oxford Health NHS Foundation Trust) for their contribution to study discussions and care of the patients included in the study.

## Footnotes

### Contributors

All listed authors certify they contributed sufficiently to this work to meet the criteria for authorship. Other contributors are listed under acknowledgements. AA, OC, AB designed the study. AA, OC, AB and IT collected the data. OC, AA and BS analysed the data. OC, AA and BS wrote the initial draft of the manuscript, OC and AA wrote the subsequent versions. All authors contributed to critical revisions of the manuscript.

## Summary box

### is already known about the subject?

Electrolyte abnormalities as part of refeeding syndrome are a well known and potentially fatal complication of weight restoration in patients with eating disorders. Existing national guidelines recommend a cautious refeeding regimen of 5-10kcal/kg/day increasing to 15-20kcal/kg/day in specialist units to reduce rates of refeeding syndrome.

### What are the new findings?

This retrospective study analyses the risk of refeeding syndrome in a large sample of severe anorexia patients who are commenced on a mean caloric regime of 26.7 kcal/day. This retrospective study shows that the vast majority of electrolyte abnormalities are mild and can be managed conservatively with oral supplementation.

### How might it impact on clinical practice in the foreseeable future?

The findings open the debate on whether the current guidelines for nutritional regimes are too cautious and may be contributing to poorer outcomes in inpatients with eating disorders. This study hopes to encourage further research into this topical area and support reconsideration of current practice.

